# Age as a Leading Indicator: A Multi-Outcome Analysis of Influenza Lead-Lag Relationships

**DOI:** 10.64898/2026.09.29.26364022

**Authors:** Emily A. Serman, Amanda Bleichrodt, Iromi Jayawardena, Carolina Liskey, Lior Rennert

## Abstract

Prior work has shown that school-age children’s influenza activity precedes that of adults by one to two weeks. Whether this is robust across outcomes or constant across a season’s course is unexamined.

Using electronic health record data from two health systems in South Carolina (2016–2026) on individuals with an influenza diagnosis or positive test, we tested age-group lead-lag relationships across six measures using differenced cross-correlation and Granger causality, with significance confirmed via heteroskedasticity-robust block permutation. We tested whether relationships differed between a season’s growth and decline phases.

Children 5–17 years led every other age group by approximately one week in four out of six outcomes and were the only group showing any nonzero-lag relationship across 60 pairwise combinations. A screen of 150 source-target combinations identified 26 permutation-confirmed instances of children’s lower-acuity activity preceding elevated emergency department or inpatient burden in other age groups. The same-outcome lead was concentrated during a season’s growth phase, while the cross-severity lead was robust throughout both a season’s growth and decline.

School-age children’s influenza activity is a robust, multi-outcome leading indicator with distinct transmission-timing and severity-progression signals in different phases of seasonal epidemics, supporting incorporation of age-stratified, phase-aware indicators into influenza forecasting.

## Introduction

Seasonal influenza epidemics impose a substantial and recurring burden on healthcare systems in the United States (US). Between 2010 and 2025, the Centers for Disease Control and Prevention (CDC) estimated that influenza caused between 9.4 and 51 million illnesses, 120,000 and 710,000 hospitalizations, and 6,300 and 52,000 deaths annually.^1^ Although the overall burden is sizeable, its heterogeneity across age, outcomes, and seasons is often obscured when examining aggregated surveillance trends. For example, during the 2024-2025 season, 57% of reported flu-related hospitalizations were attributed to those 65 years of age and older.^2^

In addition to differing in overall burden, age groups also differ in the timing of their influenza activity within a season. School-age children have repeatedly been shown to experience symptomatic illness earlier in a season than adults, plausibly reflecting the dense contact networks characteristic of school settings.^3–8^ Using emergency department syndromic surveillance in New York City, Olson et al. found that visits among children preceded the citywide rise in influenza viral isolates by approximately one week, while visits among adults 65 and older lagged the same signal.^3^ Similar age-based leads have been reported using laboratory-confirmed influenza in Utah,^7^ national hospitalization data identifying children 5-17 years as playing an outsized role in propagating the largest US influenza epidemics,^5^ and pediatric-specific surveillance identifying which age subgroups show the greatest lead time relative to population mortality.^6^ At least one prior US operational forecasting effort has identified age-stratified modeling as a promising but unimplemented direction;^9^ to our knowledge, whether and how age-specific activity might serve this purpose has not been systematically characterized.

Existing studies of age-specific lead-lag relationships in influenza share two important limitations. First, they have relied on a single surveillance outcome, typically emergency department visits or laboratory-confirmed cases, leaving it unclear whether an age-based leading indicator generalizes across the range of outcomes routinely captured by modern surveillance systems, including outpatient visits, hospitalizations, and encounter counts. Second, prior analyses have not examined whether age-specific lead-lag relationships are constant across the course of a season, despite biological plausibility that a relationship driven by active transmission (e.g., school-based spread early in a season’s growth) might differ in mechanism, and therefore in timing, from a relationship reflecting clinical progression from mild to severe illness later in a season’s course.

In this study, we used eight years of influenza surveillance data spanning six outcome measures to test whether school-age children’s influenza activity consistently leads other age groups. These outcomes captured influenza activity in two distinct ways: three measures reflecting how a case was ascertained (diagnoses, positive laboratory tests, and encounters), and three measures reflecting the clinical setting in which care was sought (outpatient, emergency department, and inpatient visits). This structure allowed us to test not only whether the same-outcome lead was robust to case-ascertainment method, but also whether school-age children’s lower-acuity activity predicted more severe outcomes in other age groups, and whether the strength and direction of these relationships differed between a season’s growth and decline phases. This work is intended to characterize the structure and consistency of these relationships as groundwork for their potential future use as forecasting inputs, rather than to develop or validate a forecasting model itself.

## Methods

### a. Data Source and Case Definition

We used aggregated electronic health record (EHR) data from the two largest health systems in South Carolina, capturing weekly influenza-related activity by patient age group from September 21, 2016 through July 28, 2026. Influenza cases were identified using International Classification of Diseases, Tenth Revision (ICD-10) J09-J11 diagnosis codes and positive influenza test results. Data were aggregated across health systems and stratified into five age groups (0–4, 5–17, 18– 49, 50–64, and 65+ years), consistent with the age stratification used by the CDC’s Influenza Hospitalization Surveillance Network (FluSurv-NET).^10^

### b. Outcome Measures

Six weekly outcome measures were examined, reflecting two distinct dimensions of influenza activity. Three measures reflected case-ascertainment method: diagnoses (based on ICD-10 coding), positive laboratory tests, and encounters (the unique union of diagnoses and positive tests occurring in a given week, avoiding double-counting of patients captured by both). Three additional measures reflected the clinical setting in which care was sought, representing increasing severity: outpatient visits, emergency department (ED) visits, and inpatient hospitalizations.

### c. Season Definition and Exclusions

Influenza seasons were defined as running from August through July of the following year, consistent with typical influenza seasonality in the continental United States. The 2020–2021 and 2021–2022 seasons were excluded from analysis due to substantially disrupted transmission dynamics associated with the COVID-19 pandemic and associated non-pharmaceutical interventions, which are not representative of typical seasonal influenza activity.^11^ The final analytic dataset comprised eight influenza seasons.

### d. Statistical Analysis

#### Differencing

For each outcome and age group, weekly counts were first-differenced (week-over-week change) prior to analysis to remove the shared seasonal trend common to all age groups, which would otherwise induce spurious contemporaneous correlation unrelated to any genuine lead-lag relationship. Stationarity of the resulting differenced series was confirmed using the augmented Dickey-Fuller test^12^ for all age groups and outcomes.

#### In-Season Restriction

To assess the robustness of the lead-lag analyses described below to the inclusion of low-activity off-season weeks, all primary analyses were additionally repeated restricted to “in-season” weeks, defined as weeks in which the statewide total for a given outcome (summed across all age groups) was at least 5% of that season’s statewide peak value for that outcome. For same-outcome analyses, this restriction was applied using the statewide total of the outcome under analysis. For cross-outcome analyses, in which a source outcome in one age group was compared to a distinct target outcome in another age group, the restriction was applied using the statewide total of the source outcome, since the analysis explicitly tests whether the target outcome’s activity is delayed relative to the source. Results are reported as “pooled” (all weeks) and “in-season” (restricted) throughout.

#### Cross-Correlation and Granger Causality

For each outcome, we computed the cross-correlation function (CCF) between every pairwise combination of age groups across lags of ±4 weeks to identify the lag at which correlation peaked. We additionally tested for Granger causality,^13^ using an autoregressive order of 4 weeks, to assess whether children 5–17 years’ activity improved prediction of other age groups’ activity beyond each group’s own autoregressive history.

#### Heteroskedasticity and Block Permutation

Diagnostic testing (multivariate autoregressive conditional heteroskedasticity [ARCH] and portmanteau tests on a fitted vector autoregression) revealed significant residual heteroskedasticity and autocorrelation in the differenced series, consistent with known volatility clustering in epidemic time series around seasonal peaks. This violates the distributional assumptions underlying the standard Granger causality F-test and can produce spuriously inflated significance; related concerns regarding the use of sample cross-correlation for identifying leading indicators in disease surveillance data have been previously described.^14^ To obtain an assumption-free estimate of significance, we implemented a block permutation procedure:^15^ the predictor series was divided into contiguous 6-week blocks, block order was randomly shuffled to preserve local volatility structure while destroying any true cross-series temporal alignment, and the Granger test statistic was recomputed across 2,000 permutations per pair to construct an empirical null distribution. Empirical p-values were calculated as the proportion of permuted statistics at least as extreme as the observed statistic.

#### Full Pairwise and Cross-Outcome Screening

We conducted a screen of all 10 pairwise age-group combinations across all six outcomes (60 total combinations) using the CCF and Granger causality approach described above. We additionally screened 150 cross-outcome, cross-age-group combinations (source age group and outcome predicting a distinct target age group and outcome, restricted to emergency department and inpatient hospitalization as target outcomes) to test whether lower-acuity activity in one age group predicted elevated higher-acuity activity in other age groups. Candidate cross-outcome pairs surviving Benjamini-Hochberg-corrected significance in this initial screen were subsequently confirmed using block permutation, evaluated in both the pooled and in-season analyses (as defined above).

#### Phase-Stratified Analysis

To test whether lead-lag relationships were constant across a season’s course, in-season weeks (as defined above) were further classified as belonging to a season’s growth (“upswing”) or decline (“downswing”) phase, based on the week-over-week direction of change in the statewide total for each outcome. For cross-outcome analyses, phase was defined using the statewide total of the source outcome, consistent with the in-season restriction described above. The CCF, Granger causality, and block permutation analyses described above were repeated separately within each phase.

#### Season-Level Timing and Magnitude Analysis

As a supplementary check on whether the week-to-week relationships described above extend to season-level prediction, we additionally tested whether children 5–17 years’ activity predicted the timing or eventual size of a season. Season onset was defined as the first week in which an age group’s activity reached a specified percentage of that season’s eventual peak value for that age group, swept across threshold values of 5%, 10%, 15%, 20%, and 25% to assess sensitivity to this choice. Peak timing was defined as the week of maximum weekly activity, and half-max timing as the first week reaching 50% of the season’s peak value, each assessed both relative to the statewide total and relative to a “rest-of-state” total excluding children 5–17, to avoid circularity from children 5–17’s contribution to the statewide total. Peak magnitude was assessed by correlating children 5–17’s cumulative activity in the weeks immediately following season onset (swept across window lengths of 2–6 weeks and the same range of onset thresholds) with that season’s eventual statewide peak value, using Spearman’s rank correlation. Because early activity and peak magnitude both increased across the study period, a partial correlation controlling for season order was additionally calculated to assess whether this association was independent of a secular time trend. All season-level analyses were limited to eight seasons (n = 8) and are reported descriptively given limited statistical power at this level of aggregation.

All analyses were conducted in R version 4.5.1.

## Results

### a. Data Characteristics

The analytic dataset comprised 410 weeks of influenza surveillance data (Supplementary Table S1) across eight influenza seasons (2016–17 through 2025–26), with the 2020–21 and 2021–22 seasons excluded due to pandemic-related disruption of typical seasonal transmission dynamics. Summary statistics for each outcome by age group are shown in Table 1, and season-by-season counts by outcome and age group are provided in Supplementary Table S2. Following first-differencing, all age-group series were confirmed stationary by augmented Dickey-Fuller testing (p < 0.01 for all age groups across all six outcomes).

**Table 1.** Case counts and proportions by age group and outcome, and proportion of weeks with zero cases (Diagnoses, Positive Tests, Encounters, Outpatient, ED Visits, and Inpatient Hospitalizations), 2016–17 through 2025–26 influenza seasons (excluding 2020–21 and 2021–22).

| Row | 0-4 | 5-17 | 18-49 | 50-64 | 65+ | Total |
| --- | --- | --- | --- | --- | --- | --- |
| Diagnoses,<br>n (%) | 38,643 (16.3) | 86,825 (36.5) | 65,274 (27.5) | 23,804 (10.0) | 23,077<br>(9.7) | 237,623 |
| Positive Tests,<br>n (%) | 33,669 (16.8) | 75,366 (37.5) | 53,479 (26.6) | 19,241<br>(9.6) | 18,966<br>(9.4) | 200,721 |
| Encounters,<br>n (%) | 43,290 (16.3) | 95,703 (36.1) | 72,969 (27.5) | 26,840 (10.1) | 26,454 (10.0) | 265,256 |
| Outpatient,<br>n (%) | 22,082 (17.0) | 56,876 (43.7) | 28,522 (21.9) | 11,635<br>(8.9) | 10,987<br>(8.4) | 130,102 |
| ED Visits,<br>n (%) | 15,384 (16.3) | 28,637 (30.3) | 34,581 (36.5) | 9,561 (10.1) | 6,467<br>(6.8) | 94,630 |
| Inpatient<br>Hospitalizations,<br>n (%) | 1,177<br>(9.1) | 1,312 (10.2) | 2,171 (16.8) | 2,608 (20.2) | 5,623 (43.6) | 12,891 |
| % weeks with<br>zero cases,<br>Encounters | 7.1 | 7.1 | 1.5 | 4.6 | 4.6 | — |
| % weeks with<br>zero cases,<br>Inpatient<br>Hospitalizations | 42.2 | 44.4 | 32.2 | 33.4 | 21 | — |

### b. Same-Outcome Age-Group Lead-Lag Relationships

Across four of six outcomes (diagnoses, emergency department visits, encounters, and positive laboratory tests), children 5–17 years showed a consistent lead of approximately one week relative to every other age group, with peak cross-correlation coefficients ranging from 0.58 to 0.75 (Table 2, Figure 1A-C, E). Outpatient visits showed a partial pattern, with children 5–17 leading only adults 65+ (r = 0.586) and showing simultaneous (lag = 0) movement relative to the remaining three age groups. This lead was confirmed using block permutation testing (2,000 permutations per pair): 22 of 24 tested age-group/outcome combinations returned an empirical p-value of 0 (more extreme than every permuted null), and the remaining two combinations (outpatient versus adults 50–64, and inpatient hospitalization versus adults 50–64) were significant at empirical p = 0.002 (Table 2).

**Figure 1:**
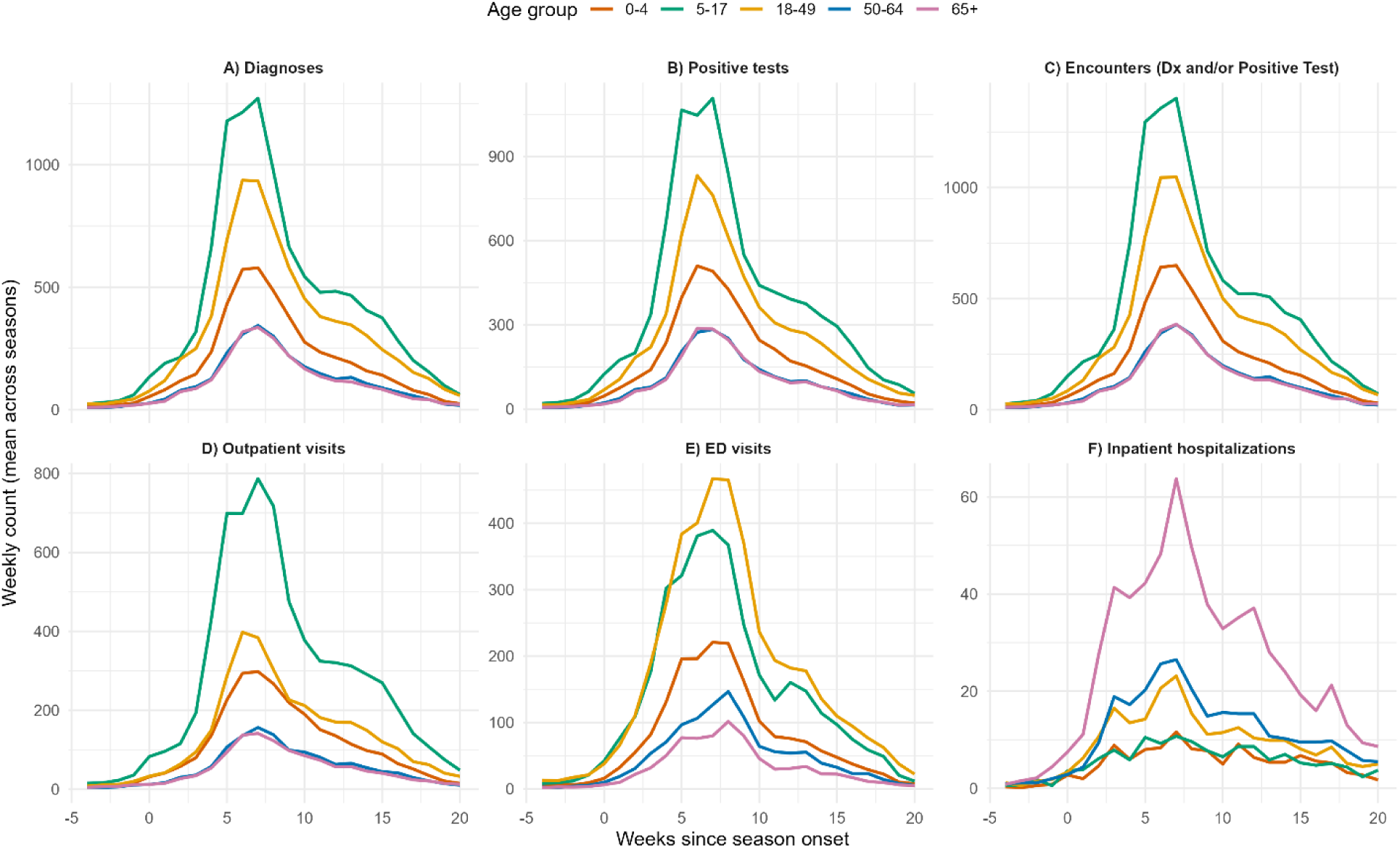
Season-averaged weekly influenza activity by age group and outcome, aligned on season onset. For each outcome, season onset was defined as the first week in which the statewide weekly total reached 5% of that season’s eventual statewide peak; curves for each age group were then aligned to their respective season’s onset week (week 0) and averaged across 8 seasons (pandemic-disrupted seasons 2020–21 and 2021–22 excluded). Panels show, from top left: (A) diagnoses, (B) positive tests, (C) encounters (diagnosis and/or positive test), (D) outpatient visits, (E) ED visits, and (F) inpatient hospitalizations. Lines represent the mean weekly count across seasons for each age group (0–4, 5–17, 18–49, 50–64, 65+ years); y-axis scales vary by panel.

**Table 2.** Same-outcome age-group lead-lag relationships: children 5–17 years versus each other age group, pooled (all weeks) and restricted to in-season weeks (. ≥**5% of that season’s peak). Positive lead counts and cross-correlation coefficients (r) reflect children 5–17 leading; empirical p-values are from block permutation testing (2,000 permutations per pair).**

| **Outcome** | **Other group** | **No. of weeks 5-17 leads (Pooled)** | **No. of weeks 5-17 leads (In season)** | **r (Pooled)** | **r (In season)** | **Empirical p (Pooled)** | **Empirical p (In season)** |
| --- | --- | --- | --- | --- | --- | --- | --- |
| Diagnoses | 0-4 | 1 | 1 | 0.726 | 0.723 | 0 | 0 |
| Diagnoses | 18-49 | 1 | 1 | 0.673 | 0.668 | 0 | 0 |
| Diagnoses | 50-64 | 1 | 1 | 0.59 | 0.585 | 0 | 0 |
| Diagnoses | 65+ | 1 | 1 | 0.597 | 0.596 | 0 | 0 |
| Positive Tests | 0-4 | 1 | 1 | 0.748 | 0.746 | 0 | 0 |
| Positive Tests | 18-49 | 1 | 1 | 0.704 | 0.702 | 0 | 0 |
| Positive Tests | 50-64 | 1 | 1 | 0.633 | 0.63 | 0 | 0 |
| Positive Tests | 65+ | 1 | 1 | 0.649 | 0.649 | 0 | 0 |
| Encounters | 0-4 | 1 | 1 | 0.737 | 0.733 | 0 | 0 |
| Encounters | 18-49 | 1 | 1 | 0.691 | 0.686 | 0 | 0 |
| Encounters | 50-64 | 1 | 1 | 0.607 | 0.603 | 0 | 0 |
| Encounters | 65+ | 1 | 1 | 0.612 | 0.611 | 0 | 0 |
| Outpatient | 0-4 | 0 | 0 | 0.712 | 0.712 | 0 | 0 |
| Outpatient | 18-49 | 0 | 0 | 0.578 | 0.577 | 0 | 0.002 |
| Outpatient | 50-64 | 0 | 0 | 0.58 | 0.581 | 0.002 | 0.01 |
| Outpatient | 65+ | 1 | 1 | 0.586 | 0.589 | 0 | 0 |
| ED Visits | 0-4 | 1 | 1 | 0.69 | 0.683 | 0 | 0 |
| ED Visits | 18-49 | 1 | 1 | 0.713 | 0.705 | 0 | 0 |
| ED Visits | 50-64 | 1 | 1 | 0.585 | 0.582 | 0 | 0 |
| ED Visits | 65+ | 1 | 1 | 0.597 | 0.596 | 0 | 0 |
| Inpatient Hospitalizations | 0-4 | 0 | 0 | 0.456 | 0.463 | 0 | 0 |
| Inpatient Hospitalizations | 18-49 | 0 | 0 | 0.474 | 0.484 | 0 | 0.001 |
| Inpatient Hospitalizations | 50-64 | 0 | 0 | 0.509 | 0.541 | 0.002 | 0.05 |
| Inpatient Hospitalizations | 65+ | 0 | 0 | 0.527 | 0.549 | 0 | 0.01 |

Inpatient hospitalization was the only outcome in which children 5–17 did not show a one-directional lead; all four age-group comparisons showed simultaneous (lag = 0) movement rather than a detectable lag in either direction (Table 2, Figure 1F). This pattern is consistent with the substantially lower baseline hospitalization rate among children relative to other age groups (Table 1), which may limit the resolution of a directional signal for this specific outcome.

### c. Full Pairwise Screen

To confirm that this lead-lag structure was not an artifact of restricting analysis to age-group pairs suggested by prior literature, we examined all 10 possible pairwise age-group combinations across all six outcomes (60 total comparisons). Across all 60 comparisons, every pair excluding children 5–17 peaked at lag 0, while children 5–17 were the only age group showing a consistent, nonzero-lag relationship with any other age group (Supplementary Table S3).

### d. Cross-Outcome, Cross-Severity Relationships

An unbiased screen of 150 source-target combinations (three lower-acuity source outcomes for children 5–17 [diagnoses, outpatient visits, positive tests] against emergency department and inpatient hospitalization outcomes in every age group) identified 26 combinations with a nonzero lag and Benjamini-Hochberg-corrected significance in the initial screen. All 26 were subsequently confirmed by block permutation testing in the pooled analysis (empirical p = 0 for all pairs, 2,000 permutations each; Table 3). Restriction to in-season weeks confirmed 23 of these 26 combinations at empirical p = 0, with the remaining three significant at empirical p ≤ 0.002 (Table 3). In each case, children 5–17’s lower-acuity activity in a given week preceded elevated emergency department or inpatient burden in other age groups by one week. The same screen also identified five candidate combinations in which another age group’s activity preceded children 5–17’s emergency department visits; two of these five reached Benjamini-Hochberg-corrected significance in the pooled analysis, but none remained significant after restriction to in-season weeks (empirical p = 0.15–0.29; Table 3) and are not interpreted as robust findings.

**Table 3.** Cross-outcome, cross-severity screen: children 5–17 years’ lower-acuity activity (diagnoses, outpatient visits, positive tests) predicting emergency department or inpatient burden in other age groups. The final five rows show the reverse direction (other age group as source) for comparison. Lag is in weeks (negative = children 5–17 lead); empirical p-values are from block permutation testing.

| Age group A | Outcome A | Age group B | Outcome B | Direction | Lag (pooled) | r (pooled) | Emp. p (pooled) | Lag (in-season) | r (in-season) | Emp. p (in-season) |
| --- | --- | --- | --- | --- | --- | --- | --- | --- | --- | --- |
| 5-17 | Positive Tests | 5-17 | Inpatient Hospitalizations | 5-17 as source | -1 | 0.5248 | 0 | -1 | 0.5357 | 0 |
| 5-17 | Positive Tests | 0-4 | ED Visits | 5-17 as source | -1 | 0.7558 | 0 | -1 | 0.7539 | 0 |
| 5-17 | Diagnoses | 0-4 | ED Visits | 5-17 as source | -1 | 0.7242 | 0 | -1 | 0.7212 | 0 |
| 5-17 | Outpatient | 5-17 | Inpatient Hospitalizations | 5-17 as source | -1 | 0.5079 | 0 | -1 | 0.5151 | 0 |
| 5-17 | Outpatient | 0-4 | ED Visits | 5-17 as source | -1 | 0.7059 | 0 | -1 | 0.7031 | 0 |
| 5-17 | Positive Tests | 18-49 | ED Visits | 5-17 as source | -1 | 0.7183 | 0 | -1 | 0.7139 | 0 |
| 5-17 | Positive Tests | 65+ | ED Visits | 5-17 as source | -1 | 0.6693 | 0 | -1 | 0.6683 | 0 |
| 5-17 | Diagnoses | 18-49 | ED Visits | 5-17 as source | -1 | 0.6937 | 0 | -1 | 0.6871 | 0 |
| 5-17 | Positive Tests | 0-4 | Inpatient Hospitalizations | 5-17 as source | -1 | 0.3849 | 0 | -1 | 0.3985 | 0 |
| 5-17 | Diagnoses | 65+ | ED Visits | 5-17 as source | -1 | 0.6344 | 0 | -1 | 0.6321 | 0 |
| 5-17 | Outpatient | 65+ | ED Visits | 5-17 as source | -1 | 0.6195 | 0 | -1 | 0.6173 | 0 |
| 5-17 | Outpatient | 18-49 | ED Visits | 5-17 as source | -1 | 0.6464 | 0 | -1 | 0.6403 | 0 |
| 5-17 | Positive Tests | 18-49 | Inpatient Hospitalizations | 5-17 as source | -1 | 0.5243 | 0 | -1 | 0.5266 | 0 |
| 5-17 | Positive Tests | 50-64 | ED Visits | 5-17 as source | -1 | 0.6259 | 0 | -1 | 0.6227 | 0 |
| 5-17 | Outpatient | 0-4 | Inpatient Hospitalizations | 5-17 as source | -1 | 0.3548 | 0 | -1 | 0.3655 | 0 |
| 5-17 | Diagnoses | 0-4 | Inpatient Hospitalizations | 5-17 as source | -1 | 0.3313 | 0 | -1 | 0.3415 | 0 |
| 5-17 | Diagnoses | 50-64 | ED Visits | 5-17 as source | -1 | 0.5962 | 0 | -1 | 0.5912 | 0 |
| 5-17 | Diagnoses | 18-49 | Inpatient Hospitalizations | 5-17 as source | -1 | 0.4855 | 0 | -1 | 0.4871 | 0 |
| 5-17 | Outpatient | 50-64 | ED Visits | 5-17 as source | -1 | 0.5707 | 0 | -1 | 0.566 | 0 |
| 5-17 | Positive Tests | 50-64 | Inpatient Hospitalizations | 5-17 as source | -1 | 0.5385 | 0 | -1 | 0.5435 | 0 |
| 5-17 | Outpatient | 18-49 | Inpatient Hospitalizations | 5-17 as source | -1 | 0.4579 | 0 | -1 | 0.4583 | 0 |
| 5-17 | Outpatient | 50-64 | Inpatient Hospitalizations | 5-17 as source | -1 | 0.5025 | 0 | -1 | 0.5035 | 0.001 |
| 5-17 | Diagnoses | 50-64 | Inpatient Hospitalizations | 5-17 as source | -1 | 0.5017 | 0 | -1 | 0.5044 | 0 |
| 5-17 | Positive Tests | 65+ | Inpatient Hospitalizations | 5-17 as source | -1 | 0.5396 | 0 | -1 | 0.5411 | 0 |
| 5-17 | Outpatient | 65+ | Inpatient Hospitalizations | 5-17 as source | -1 | 0.4997 | 0 | -1 | 0.5 | <0.001 |
| 5-17 | Diagnoses | 65+ | Inpatient Hospitalizations | 5-17 as source | -1 | 0.5003 | 0 | -1 | 0.5009 | 0.002 |
| 50-64 | Positive Tests | 5-17 | ED Visits | other age group as source | 1 | 0.548 | 0.02 | 1 | 0.5458 | 0.15 |
| 50-64 | Diagnoses | 5-17 | ED Visits | other age group as source | 1 | 0.5413 | 0.03 | 1 | 0.5365 | 0.17 |
| 65+ | Outpatient | 5-17 | ED Visits | other age group as source | 1 | 0.4106 | 0.05 | 1 | 0.4129 | 0.16 |
| 65+ | Diagnoses | 5-17 | ED Visits | other age group as<br>source | 1 | 0.5072 | 0.06 | 1 | 0.5055 | 0.29 |
| 65+ | Positive Tests | 5-17 | ED Visits | other age<br>group as<br>source | 1 | 0.5222 | 0.07 | 1 | 0.5228 | 0.27 |

### e. Robustness to Off-Season Weeks

Restricting the same-outcome analysis to in-season weeks only (weeks with total influenza activity ≥5% of that season’s peak, to exclude the low-activity off-season floor) produced results nearly identical to the full, unrestricted analysis: lag direction was unchanged for all 24 tested age-group/outcome combinations, correlation coefficients differed by less than 0.05 in all cases, and all combinations remained significant under block permutation (empirical p ≤ 0.049; Table 2).

### f. Phase-Stratified Analysis

Weeks were classified as belonging to a season’s growth (“upswing”) or decline (“downswing”) phase, restricted to in-season weeks as defined above. Following block permutation confirmation in both pooled and in-season analyses, the same-outcome lead was concentrated during a season’s growth phase for adults 18–49, with 5 of 6 outcomes confirmed robust during the upswing compared to 1 of 6 during the downswing (Table 4). This upswing-concentrated pattern was weaker for adults 50–64 (2 of 6 outcomes robust during the upswing, 0 of 6 during the downswing) and was absent for adults 65+, where the lead was instead present to a similar, modest degree in both phases (2 of 6 outcomes robust in each). In contrast, the relationship with children 0–4 was concentrated during the downswing (3 of 6 outcomes robust) rather than the upswing (0 of 6 robust).

**Table 4.** Phase-stratified same-outcome lead-lag relationships between children 5–17 years and each other age group, by season phase (upswing = growth, downswing = decline).

| Outcome | Comparison group | Phase | Weeks 5-17 leads (pooled) | r (pooled) | Weeks 5-17 leads (in-season) | r (in-season) | Emp. p (pooled) | Emp. p (in-season) |
| --- | --- | --- | --- | --- | --- | --- | --- | --- |
| Diagnoses | 0-4 | downswing | 1 | 0.5605 | 0 | 0.4153 | 0.001 | 0.10 |
| Encounters | 0-4 | downswing | 1 | 0.5845 | 1 | 0.4484 | 0.001 | 0.07 |
| Inpatient Hospitalizations | 0-4 | downswing | 1 | 0.376 | 1 | 0.3146 | 0.002 | 0.03 |
| Outpatient | 0-4 | downswing | 1 | 0.6408 | 1 | 0.5154 | 0 | 0.004 |
| Positive Tests | 0-4 | downswing | 1 | 0.6357 | 1 | 0.5218 | 0 | 0.002 |
| ED Visits | 0-4 | upswing | 1 | 0.5305 | 1 | 0.4318 | 0.001 | 0.08 |
| Positive Tests | 0-4 | upswing | 1 | 0.5825 | 1 | 0.4967 | 0.003 | 0.06 |
| Outpatient | 18-49 | downswing | 1 | 0.5747 | 1 | 0.5037 | 0 | 0.005 |
| Diagnoses | 18-49 | upswing | 1 | 0.5563 | 1 | 0.4426 | <0.001 | 0.04 |
| ED Visits | 18-49 | upswing | 1 | 0.6778 | 1 | 0.6204 | 0 | 0 |
| Encounters | 18-49 | upswing | 1 | 0.5908 | 1 | 0.4841 | 0 | 0.02 |
| Inpatient Hospitalizations | 18-49 | upswing | 1 | 0.4458 | 1 | 0.4591 | 0 | 0.04 |
| Positive Tests | 18-49 | upswing | 1 | 0.572 | 1 | 0.4925 | 0.002 | <0.001 |
| Outpatient | 50-64 | downswing | 1 | 0.4559 | 1 | 0.3525 | 0.002 | 0.06 |
| Diagnoses | 50-64 | upswing | 1 | 0.4832 | 1 | 0.3486 | 0.001 | 0.13 |
| ED Visits | 50-64 | upswing | 1 | 0.5836 | 1 | 0.5013 | 0 | 0 |
| Encounters | 50-64 | upswing | 1 | 0.5273 | 1 | 0.4033 | 0 | 0.06 |
| Inpatient Hospitalizations | 50-64 | upswing | 0 | 0.3472 | -2 | 0.4243 | 0.008 | 0.35 |
| Positive Tests | 50-64 | upswing | 1 | 0.5237 | 1 | 0.4224 | 0 | 0.001 |
| Diagnoses | 65+ | downswing | 1 | 0.3777 | 1 | 0.2509 | 0.07 | 0.54 |
| ED Visits | 65+ | downswing | 1 | 0.54 | 1 | 0.4356 | 0.001 | 0.01 |
| Encounters | 65+ | downswing | 1 | 0.3855 | 1 | 0.2703 | 0.06 | 0.48 |
| Outpatient | 65+ | downswing | 1 | 0.4949 | 1 | 0.4492 | 0 | 0.04 |
| Diagnoses | 65+ | upswing | 1 | 0.4753 | 1 | 0.3648 | 0.001 | 0.11 |
| ED Visits | 65+ | upswing | 1 | 0.5223 | 1 | 0.4352 | 0 | 0 |
| Encounters | 65+ | upswing | 1 | 0.5076 | 1 | 0.4032 | 0.002 | 0.07 |
| Positive Tests | 65+ | upswing | 1 | 0.531 | 1 | 0.4514 | <0.001 | <0.001 |

In contrast, the cross-outcome relationship described above was present throughout the season rather than concentrated in a single phase. Across 36 candidate pair-phase combinations screened from the 26 confirmed cross-outcome pairs (Table 3), block permutation testing confirmed 12 of 17 upswing-phase candidates (71%) and 11 of 19 downswing-phase candidates (58%) as robust in both pooled and in-season analyses (Table 5). This indicates the cross-severity lead reflects a comparatively stable, approximately one-week relationship that persists across a season’s rise and decline, distinct from the upswing-concentrated same-outcome lead described above.

**Table 5.** Phase-stratified cross-outcome, cross-severity relationships between children 5–17 years’ lower-acuity activity and emergency department/inpatient burden in other age groups, by season phase.

| Age group A | Outcome A | Age group B | Outcome B | Phase | Lag (pooled) | r (pooled) | Lag (in-season) | r (in-season) | Emp. p (pooled) | Emp. p (in-season) |
| --- | --- | --- | --- | --- | --- | --- | --- | --- | --- | --- |
| 5-17 | Positive Tests | 0-4 | ED Visits | downswing | -1 | 0.59 | -1 | 0.4893 | 0 | <0.001 |
| 5-17 | Diagnoses | 0-4 | ED Visits | downswing | -1 | 0.5031 | -1 | 0.3651 | 0 | 0.15 |
| 5-17 | Outpatient | 5-17 | Inpatient Hospitalizations | downswing | -1 | 0.568 | -1 | 0.5361 | 0 | 0 |
| 5-17 | Outpatient | 0-4 | ED Visits | downswing | -1 | 0.6066 | -1 | 0.5478 | 0 | 0.003 |
| 5-17 | Positive Tests | 18-49 | ED Visits | downswing | -1 | 0.595 | -1 | 0.4754 | 0 | 0 |
| 5-17 | Positive Tests | 65+ | ED Visits | downswing | -1 | 0.52 | -1 | 0.4283 | 0 | 0.002 |
| 5-17 | Diagnoses | 18-49 | ED Visits | downswing | -1 | 0.5316 | -1 | 0.3963 | 0 | 0.07 |
| 5-17 | Outpatient | 18-49 | ED Visits | downswing | -1 | 0.5679 | -1 | 0.495 | 0 | 0.004 |
| 5-17 | Outpatient | 0-4 | Inpatient Hospitalizations | downswing | -2 | 0.3058 | -2 | 0.2744 | 0 | 0.01 |
| 5-17 | Diagnoses | 50-64 | ED Visits | downswing | -1 | 0.451 | -1 | 0.3323 | 0 | 0.06 |
| 5-17 | Outpatient | 50-64 | ED Visits | downswing | -1 | 0.4776 | -1 | 0.413 | 0 | 0.01 |
| 5-17 | Outpatient | 18-49 | Inpatient Hospitalizations | downswing | -1 | 0.3874 | -1 | 0.3403 | 0.002 | 0.02 |
| 5-17 | Diagnoses | 50-64 | Inpatient Hospitalizations | downswing | -1 | 0.3448 | -1 | 0.2586 | 0.002 | 0.17 |
| 5-17 | Outpatient | 50-64 | Inpatient Hospitalizations | downswing | -1 | 0.3916 | -1 | 0.3594 | 0.003 | 0.02 |
| 5-17 | Diagnoses | 65+ | ED Visits | downswing | -1 | 0.4309 | -1 | 0.3302 | 0.004 | 0.14 |
| 5-17 | Outpatient | 65+ | ED Visits | downswing | -1 | 0.4603 | -1 | 0.4256 | 0.004 | 0.04 |
| 5-17 | Positive Tests | 65+ | Inpatient Hospitalizations | downswing | -1 | 0.4445 | -1 | 0.3338 | 0.005 | 0.09 |
| 5-17 | Outpatient | 65+ | Inpatient Hospitalizations | downswing | -1 | 0.3679 | -1 | 0.2901 | 0.005 | 0.08 |
| 5-17 | Diagnoses | 65+ | Inpatient Hospitalizations | downswing | -1 | 0.365 | -1 | 0.236 | 0.03 | 0.46 |
| 5-17 | Positive Tests | 0-4 | ED Visits | upswing | -1 | 0.5927 | -1 | 0.5133 | 0 | 0.05 |
| 5-17 | Diagnoses | 0-4 | ED Visits | upswing | -1 | 0.5987 | -1 | 0.5075 | 0 | 0.02 |
| 5-17 | Positive Tests | 18-49 | ED Visits | upswing | -1 | 0.5821 | -1 | 0.4922 | 0 | 0.03 |
| 5-17 | Positive Tests | 65+ | ED Visits | upswing | -1 | 0.5086 | -1 | 0.4187 | 0 | 0.03 |
| 5-17 | Diagnoses | 18-49 | ED Visits | upswing | -1 | 0.5844 | -1 | 0.4806 | 0 | 0.03 |
| 5-17 | Positive Tests | 0-4 | Inpatient Hospitalizations | upswing | -2 | 0.3985 | -2 | 0.3044 | 0 | 0.05 |
| 5-17 | Positive Tests | 18-49 | Inpatient Hospitalizations | upswing | -1 | 0.4803 | -1 | 0.4063 | 0 | 0.03 |
| 5-17 | Positive Tests | 50-64 | ED Visits | upswing | -1 | 0.5374 | -1 | 0.4425 | 0 | 0.03 |
| 5-17 | Diagnoses | 0-4 | Inpatient Hospitalizations | upswing | -2 | 0.4378 | -2 | 0.3812 | 0 | 0.06 |
| 5-17 | Diagnoses | 50-64 | ED Visits | upswing | -1 | 0.5214 | -1 | 0.3886 | 0 | 0.06 |
| 5-17 | Diagnoses | 18-49 | Inpatient Hospitalizations | upswing | -1 | 0.5146 | -1 | 0.4531 | 0 | 0.03 |
| 5-17 | Positive Tests | 50-64 | Inpatient Hospitalizations | upswing | -1 | 0.4316 | -1 | 0.3404 | 0 | 0.03 |
| 5-17 | Positive Tests | 65+ | Inpatient Hospitalizations | upswing | -1 | 0.4976 | -1 | 0.4097 | 0 | 0.04 |
| 5-17 | Diagnoses | 65+ | ED Visits | upswing | -1 | 0.4948 | -1 | 0.4021 | <0.001 | 0.03 |
| 5-17 | Positive Tests | 5-17 | Inpatient Hospitalizations | upswing | 0 | 0.3714 | -4 | -0.3662 | 0.002 | 0.05 |
| 5-17 | Diagnoses | 65+ | Inpatient Hospitalizations | upswing | -1 | 0.4252 | -2 | 0.3525 | 0.005 | 0.19 |
| 5-17 | Diagnoses | 50-64 | Inpatient Hospitalizations | upswing | -1 | 0.3783 | -1 | 0.2684 | 0.006 | 0.30 |

### g. Season-Level Timing and Magnitude

We additionally tested whether children 5–17’s activity predicted season-level onset timing, peak timing, or peak magnitude at the individual-season level (n = 8 seasons). Across multiple operationalizations of onset and peak timing, tested at several threshold values, no consistent or threshold-robust lead was observed (Supplementary Tables S4–S6). An initial bivariate correlation between early-season activity in children 5–17 and eventual peak magnitude (Spearman’s ρ = 0.81, p = 0.02) did not remain significant after controlling for a secular time trend present across the study period (partial ρ = 0.54, p = 0.21) and is not interpreted as a robust finding (Supplementary Table S7).

## Discussion

In this analysis of eight years of influenza surveillance data from two large healthcare systems, school-age children (5-17 years) showed a consistent, approximately one-week lead over every other age group across four of six outcome measures, with a partial pattern in a fifth, and were the only age group to show any nonzero-lag relationship when considering all possible age-group pairings. This lead extended beyond same-outcome comparisons: children’s lower-acuity activity predicted elevated emergency department and inpatient burden in other age groups one week later, across 26 permutation-confirmed source-target combinations. Notably, these two relationships differed in their relationship to a season’s phase: the same-outcome lead was concentrated during a season’s growth phase, particularly for adults 18–49, while the cross-severity lead was robust throughout both a season’s growth and decline.

These findings are consistent with, and extend, prior work identifying school-age children as an early signal in seasonal influenza activity. Using single-outcome data, prior studies in New York City,^3^ Utah,^4^ and Massachusetts^6^ have each reported that pediatric or school-age influenza activity precedes that of older age groups by approximately one to two weeks, and national hospitalization surveillance data have identified children 5-17 years as disproportionately contributing to the largest US influenza epidemics.^5^ The present study extends this literature in three respects. First, by testing across three case ascertainment measures and three outcome measures spanning distinct points along the clinical care-seeking pathway, we show that the same-outcome lead is robust across most outcomes, while also identifying inpatient hospitalization as the one outcome in which no directional lead was detected. Second, we identify a distinct cross-severity relationship not previously described in this literature, in which children’s lower-acuity activity predicts elevated severe-outcome burden in other age groups. Third, by testing for phase-dependence, we show that the same-outcome lead, but not the cross-severity relationship, varies across the course of a season, indicating these two relationships likely reflect distinct underlying mechanisms.

We interpret the differing phase-dependence of these two relationships as consistent with two distinct, but not mutually exclusive, mechanisms. The same-outcome lead, concentrated during a season’s growth phase, is consistent with a transmission-timing interpretation: school-based contact networks may seed early spread that subsequently propagates to other age groups as a season’s epidemic curve rises, a mechanism that would be expected to weaken once an epidemic is established and its decline is instead driven by more synchronized factors, such as depletion of susceptible individuals or behavioral change, across all age groups simultaneously. The cross-severity lead, by contrast, held consistently across both a season’s growth and decline, which is more consistent with a stable, within-person clinical-progression interval between an initial lower-acuity presentation and a subsequent, more severe outcome. If children 5–17 continue to represent the leading edge of a season’s trajectory throughout its course, not only during its initial growth, a roughly constant lag between their lower-acuity activity and other age groups’ more severe outcomes would be expected regardless of whether case counts are rising or falling in a given week. We emphasize that this study did not directly test either mechanism, and this interpretation should be treated as a plausible explanation for the observed pattern rather than a demonstrated causal pathway.

Unlike the other five outcomes, inpatient hospitalization showed no detectable directional lead between children 5-17 and any other age group; all comparisons showed simultaneous (lag = 0) movement. This is plausibly explained by the substantially lower rate of pediatric hospitalization relative to other age groups:^16^ children accounted for the fewest inpatient encounters and had the highest proportion of weeks with zero hospitalizations of any age group in this dataset (Table 1). A sparser, less complete signal in this specific outcome and age group may limit the ability to detect a clean, one-directional lead, even if the same underlying transmission-timing relationship is present.

Despite the robust week-to-week lead described above, children’s activity did not reliably predict season-level onset timing, peak timing, or peak magnitude when tested at the level of individual seasons (n = 8). We interpret this as reflecting a meaningful distinction between two related but different phenomena: a consistent relationship in the ongoing, week-to-week rhythm of a season once it is underway, versus a reliable predictor of when a season will begin or how large it will ultimately become. Season onset may be more strongly influenced by factors not captured in this analysis, such as the timing of introduction or importation of circulating strains, than by age-specific transmission dynamics within the state. This distinction has direct relevance for forecasting applications: the relationships identified here may be better suited to short-term, within-season indicators than to early prediction of a season’s overall trajectory.

Accurate forecasts of influenza onset, peak timing, and severity can assist with staffing, resource allocation, and public communication. The consistency of the relationship identified in this analysis across multiple outcome measures, together with their permutation-confirmed statistical robustness, suggests that age-stratified indicators, particularly children’s lower-acuity care-seeking activity as a potential short-term predictor of severe-outcome burden in other age groups, may warrant evaluation as candidate inputs in future influenza forecasting work.^9,17^

This study has several limitations. First, data were drawn from two health systems within a single state, which may limit generalizability to other regions with different demographic structures, healthcare-seeking patterns, or school calendars. Second, influenza cases were identified using diagnosis codes and positive test results without stratification by circulating strain or subtype; at least one prior study has found that the age group showing the earliest activity may vary by subtype,^7^ which may partially explain variability in the relationships we observed. Third, analyses conducted at the level of individual seasons (onset timing, peak timing, and peak magnitude) were limited by a small sample of eight seasons, constraining statistical power for these specific comparisons. Fourth, this analysis was conducted using state-level aggregated data; prior work examining influenza at varying spatial scales has found that spatial heterogeneity in outbreak timing may only be detectable at sufficiently fine geographic resolution,^18^ suggesting that geographic variation in the age-based relationships identified here is an important open question not addressed by the present analysis. Future work should continue to explore whether geographical variations (eg: local population structure, school density, or community connectivity) moderate the strength or timing of the relations identified in this manuscript^18^ and if they vary by circulating influenza subtype.^7^

School-age children’s influenza activity is a robust, multi-outcome leading indicator relative to other age groups, with distinct transmission-timing and severity-progression signals operating during different phases of a season’s course. These findings support further evaluation of age-stratified, phase-aware indicators in influenza surveillance and forecasting, while highlighting that this relationship does not straightforwardly extend to predicting when a season will begin or how large it will become.

## Acknowledgements

*AI Statement:* Claude Sonnet 5 (Anthropic) was used to assist with editing manuscript text and troubleshooting statistical code. The authors reviewed, verified, and take full responsibility for the integrity, accuracy, and content of all text in the final manuscript.

## Data Availability

Data are not publicly available due to PHI and IRB restrictions.

