## Supplemental Tables for "Age as a Leading Indicator: A Multi-Outcome Analysis of Influenza Lead-Lag Relationships"

Table of Contents

**Table S1.** Season-level summary: start/end weeks, total weeks, in-season weeks (≥5% of peak), peak week, peak magnitude, and total encounters, by influenza season	2

**Table S2.** Season- and age-group-level counts for each outcome measure (diagnoses, positive tests, encounters, outpatient visits, ED visits, inpatient hospitalizations)	3

**Table S3.** Full pairwise screen of all 10 age-group combinations across all 6 outcomes (60 comparisons), showing peak cross-correlation lag and r for each pair	4-5

**Table S4.** Sensitivity of season-onset-based lead estimates to the onset threshold used to define season start (5–25% of peak), by comparison group. Cells show fraction of seasons in which children 5–17 led (numerator/8) and the median lead in weeks	6

**Table S5.** Season-level peak-timing analysis: fraction of seasons (of 8) in which children 5–17 years led the statewide total or the rest-of-state signal at peak week and half-maximum week, with median lead in weeks	6

**Table S6.** Rank of children 5–17 years' weekly activity (of 5 age groups) in the weeks leading up to the statewide peak, across 8 seasons	6

**Table S7.** Sensitivity of the bivariate correlation between early-season children 5–17 activity and eventual peak magnitude to the early-season window length and onset threshold used to define it (Spearman's ρ)	7

**Table S1. Season-level summary: start/end weeks, total weeks, in-season weeks (≥5% of peak), peak week, peak magnitude, and total encounters, by influenza season.**

| **Season** | **Start week**  (YYYY-MM-DD) | **End week**  (YYYY-MM-DD) | **Weeks (total)** | **Weeks (in-season)** | **Peak week (statewide)** | **Peak magnitude (encounters)** | **Total encounters** |
| --- | --- | --- | --- | --- | --- | --- | --- |
| 2016-17 | 2016-09-28 | 2017-07-26 | 44 | 20 | 2017-02-15 | 1,597 | 15,603 |
| 2017-18 | 2017-08-02 | 2018-07-25 | 52 | 16 | 2018-01-31 | 3,680 | 24,121 |
| 2018-19 | 2018-08-01 | 2019-07-31 | 53 | 18 | 2019-01-30 | 2,901 | 18,219 |
| 2019-20 | 2019-08-07 | 2020-07-29 | 52 | 20 | 2020-01-29 | 2,565 | 26,489 |
| 2022-23 | 2022-08-03 | 2023-07-26 | 52 | 22 | 2022-10-26 | 4,993 | 34,767 |
| 2023-24 | 2023-08-02 | 2024-07-31 | 53 | 23 | 2023-12-13 | 5,900 | 46,176 |
| 2024-25 | 2024-08-07 | 2025-07-30 | 52 | 16 | 2025-01-29 | 8,846 | 51,789 |
| 2025-26 | 2025-08-06 | 2026-07-29 | 52 | 22 | 2025-12-24 | 6,226 | 48,092 |

**Table S2. Season- and age-group-level counts for each outcome measure (diagnoses, positive tests, encounters, outpatient visits, ED visits, inpatient hospitalizations).**

| **Season** | **Age group** | **Diagnoses** | **Positive Tests** | **Encounters** | **Outpatient** | **ED Visits** | **Inpatient Hospitalizations** |
| --- | --- | --- | --- | --- | --- | --- | --- |
| 2016-17 | 0-4 | 2,499 | 2,139 | 2,632 | 1,640 | 795 | 64 |
| 2016-17 | 5-17 | 6,396 | 5,414 | 6,747 | 4,256 | 2,048 | 92 |
| 2016-17 | 18-49 | 3,480 | 2,333 | 3,662 | 970 | 2,421 | 89 |
| 2016-17 | 50-64 | 1,348 | 955 | 1,422 | 504 | 739 | 105 |
| 2016-17 | 65+ | 1,053 | 792 | 1,140 | 410 | 437 | 206 |
| 2017-18 | 0-4 | 4,047 | 3,160 | 4,263 | 2,625 | 1,314 | 108 |
| 2017-18 | 5-17 | 7,832 | 5,988 | 8,187 | 5,066 | 2,671 | 95 |
| 2017-18 | 18-49 | 6,173 | 3,727 | 6,445 | 1,846 | 4,186 | 141 |
| 2017-18 | 50-64 | 2,876 | 1,916 | 3,072 | 1,195 | 1,480 | 201 |
| 2017-18 | 65+ | 1,970 | 1,380 | 2,154 | 843 | 728 | 399 |
| 2018-19 | 0-4 | 3,304 | 2,455 | 3,470 | 2,175 | 1,038 | 91 |
| 2018-19 | 5-17 | 7,646 | 5,637 | 7,966 | 5,069 | 2,487 | 90 |
| 2018-19 | 18-49 | 3,864 | 2,160 | 4,049 | 1,289 | 2,462 | 113 |
| 2018-19 | 50-64 | 1,370 | 830 | 1,449 | 611 | 637 | 122 |
| 2018-19 | 65+ | 1,197 | 805 | 1,285 | 514 | 426 | 257 |
| 2019-20 | 0-4 | 4,914 | 3,501 | 5,099 | 2,876 | 1,930 | 108 |
| 2019-20 | 5-17 | 9,232 | 6,800 | 9,613 | 5,707 | 3,432 | 93 |
| 2019-20 | 18-49 | 7,187 | 4,014 | 7,481 | 2,180 | 4,792 | 215 |
| 2019-20 | 50-64 | 2,426 | 1,394 | 2,592 | 980 | 1,189 | 257 |
| 2019-20 | 65+ | 1,548 | 898 | 1,704 | 654 | 525 | 369 |
| 2022-23 | 0-4 | 5,148 | 4,829 | 6,067 | 2,696 | 2,269 | 183 |
| 2022-23 | 5-17 | 12,628 | 11,729 | 14,680 | 7,559 | 4,880 | 189 |
| 2022-23 | 18-49 | 6,522 | 6,113 | 8,125 | 1,820 | 4,438 | 264 |
| 2022-23 | 50-64 | 2,224 | 2,099 | 2,830 | 866 | 1,062 | 296 |
| 2022-23 | 65+ | 2,401 | 2,304 | 3,065 | 887 | 819 | 695 |
| 2023-24 | 0-4 | 5,817 | 5,867 | 7,031 | 2,942 | 2,661 | 214 |
| 2023-24 | 5-17 | 14,286 | 14,130 | 16,644 | 9,071 | 4,941 | 274 |
| 2023-24 | 18-49 | 10,878 | 10,655 | 13,082 | 4,426 | 5,972 | 480 |
| 2023-24 | 50-64 | 4,054 | 3,914 | 4,916 | 1,958 | 1,565 | 531 |
| 2023-24 | 65+ | 3,632 | 3,442 | 4,503 | 1,729 | 981 | 922 |
| 2024-25 | 0-4 | 6,969 | 6,477 | 7,965 | 3,823 | 2,938 | 208 |
| 2024-25 | 5-17 | 13,572 | 11,938 | 15,060 | 9,080 | 4,262 | 230 |
| 2024-25 | 18-49 | 13,727 | 12,094 | 15,238 | 7,374 | 5,832 | 521 |
| 2024-25 | 50-64 | 5,741 | 4,834 | 6,350 | 3,118 | 1,927 | 696 |
| 2024-25 | 65+ | 6,410 | 5,307 | 7,176 | 3,102 | 1,557 | 1751 |
| 2025-26 | 0-4 | 5,945 | 5,241 | 6,763 | 3,305 | 2,439 | 201 |
| 2025-26 | 5-17 | 15,233 | 13,730 | 16,806 | 11,068 | 3,916 | 249 |
| 2025-26 | 18-49 | 13,443 | 12,383 | 14,887 | 8,617 | 4,478 | 348 |
| 2025-26 | 50-64 | 3,765 | 3,299 | 4,209 | 2,403 | 962 | 400 |
| 2025-26 | 65+ | 4,866 | 4,038 | 5,427 | 2,848 | 994 | 1,024 |

**Table S3. Full pairwise screen of all 10 age-group combinations across all 6 outcomes (60 comparisons), showing peak cross-correlation lag and r for each pair.**

| **Outcome** | **Age-group pair** | **Lag (weeks)** | **r** | **Involves 5-17** |
| --- | --- | --- | --- | --- |
| Diagnoses | 0-4 vs. 5-17 | 1 | 0.7257 | TRUE |
| Diagnoses | 18-49 vs. 5-17 | 1 | 0.6729 | TRUE |
| Diagnoses | 5-17 vs. 50-64 | -1 | 0.5895 | TRUE |
| Diagnoses | 5-17 vs. 65+ | -1 | 0.5969 | TRUE |
| ED Visits | 0-4 vs. 5-17 | 1 | 0.6897 | TRUE |
| ED Visits | 18-49 vs. 5-17 | 1 | 0.7127 | TRUE |
| ED Visits | 5-17 vs. 50-64 | -1 | 0.5848 | TRUE |
| ED Visits | 5-17 vs. 65+ | -1 | 0.5974 | TRUE |
| Encounters | 0-4 vs. 5-17 | 1 | 0.737 | TRUE |
| Encounters | 18-49 vs. 5-17 | 1 | 0.6907 | TRUE |
| Encounters | 5-17 vs. 50-64 | -1 | 0.6074 | TRUE |
| Encounters | 5-17 vs. 65+ | -1 | 0.6119 | TRUE |
| Inpatient Hospitalizations | 0-4 vs. 5-17 | 0 | 0.4558 | TRUE |
| Inpatient Hospitalizations | 18-49 vs. 5-17 | 0 | 0.4741 | TRUE |
| Inpatient Hospitalizations | 5-17 vs. 50-64 | 0 | 0.5089 | TRUE |
| Inpatient Hospitalizations | 5-17 vs. 65+ | 0 | 0.5271 | TRUE |
| Outpatient | 0-4 vs. 5-17 | 0 | 0.7119 | TRUE |
| Outpatient | 18-49 vs. 5-17 | 0 | 0.5776 | TRUE |
| Outpatient | 5-17 vs. 50-64 | 0 | 0.5796 | TRUE |
| Outpatient | 5-17 vs. 65+ | -1 | 0.5864 | TRUE |
| Positive Tests | 0-4 vs. 5-17 | 1 | 0.7481 | TRUE |
| Positive Tests | 18-49 vs. 5-17 | 1 | 0.7042 | TRUE |
| Positive Tests | 5-17 vs. 50-64 | -1 | 0.633 | TRUE |
| Positive Tests | 5-17 vs. 65+ | -1 | 0.6492 | TRUE |
| Diagnoses | 0-4 vs. 18-49 | 0 | 0.9358 | FALSE |
| Diagnoses | 0-4 vs. 50-64 | 0 | 0.8812 | FALSE |
| Diagnoses | 0-4 vs. 65+ | 0 | 0.8397 | FALSE |
| Diagnoses | 18-49 vs. 50-64 | 0 | 0.9297 | FALSE |
| Diagnoses | 18-49 vs. 65+ | 0 | 0.8903 | FALSE |
| Diagnoses | 50-64 vs. 65+ | 0 | 0.9116 | FALSE |
| ED Visits | 0-4 vs. 18-49 | 0 | 0.9199 | FALSE |
| ED Visits | 0-4 vs. 50-64 | 0 | 0.8531 | FALSE |
| ED Visits | 0-4 vs. 65+ | 0 | 0.854 | FALSE |
| ED Visits | 18-49 vs. 50-64 | 0 | 0.9016 | FALSE |
| ED Visits | 18-49 vs. 65+ | 0 | 0.8434 | FALSE |
| ED Visits | 50-64 vs. 65+ | 0 | 0.8537 | FALSE |
| Encounters | 0-4 vs. 18-49 | 0 | 0.9458 | FALSE |
| Encounters | 0-4 vs. 50-64 | 0 | 0.8928 | FALSE |
| Encounters | 0-4 vs. 65+ | 0 | 0.8439 | FALSE |
| Encounters | 18-49 vs. 50-64 | 0 | 0.9311 | FALSE |
| Encounters | 18-49 vs. 65+ | 0 | 0.8918 | FALSE |
| Encounters | 50-64 vs. 65+ | 0 | 0.9201 | FALSE |
| Inpatient Hospitalizations | 0-4 vs. 18-49 | 0 | 0.5212 | FALSE |
| Inpatient Hospitalizations | 0-4 vs. 50-64 | 0 | 0.5124 | FALSE |
| Inpatient Hospitalizations | 0-4 vs. 65+ | 0 | 0.5498 | FALSE |
| Inpatient Hospitalizations | 18-49 vs. 50-64 | 0 | 0.6666 | FALSE |
| Inpatient Hospitalizations | 18-49 vs. 65+ | 0 | 0.6786 | FALSE |
| Inpatient Hospitalizations | 50-64 vs. 65+ | 0 | 0.7715 | FALSE |
| Outpatient | 0-4 vs. 18-49 | 0 | 0.815 | FALSE |
| Outpatient | 0-4 vs. 50-64 | 0 | 0.8068 | FALSE |
| Outpatient | 0-4 vs. 65+ | 0 | 0.7124 | FALSE |
| Outpatient | 18-49 vs. 50-64 | 0 | 0.8935 | FALSE |
| Outpatient | 18-49 vs. 65+ | 0 | 0.8787 | FALSE |
| Outpatient | 50-64 vs. 65+ | 0 | 0.8559 | FALSE |
| Positive Tests | 0-4 vs. 18-49 | 0 | 0.923 | FALSE |
| Positive Tests | 0-4 vs. 50-64 | 0 | 0.8838 | FALSE |
| Positive Tests | 0-4 vs. 65+ | 0 | 0.8398 | FALSE |
| Positive Tests | 18-49 vs. 50-64 | 0 | 0.9265 | FALSE |
| Positive Tests | 18-49 vs. 65+ | 0 | 0.9003 | FALSE |
| Positive Tests | 50-64 vs. 65+ | 0 | 0.9208 | FALSE |

**Table S4. Sensitivity of season-onset-based lead estimates to the onset threshold used to define season start (5–25% of peak), by comparison group. Cells show fraction of seasons in which children 5–17 led (numerator/8) and the median lead in weeks.**

| **Comparison group** | **5%** | **10%** | **15%** | **20%** | **25%** |
| --- | --- | --- | --- | --- | --- |
| 0-4 | 2/8 (+0 wk) | 2/8 (+0 wk) | 2/8 (-1 wk) | 1/8 (+0 wk) | 2/8 (+0 wk) |
| 18-49 | 2/8 (-0.5 wk) | 4/8 (+0.5 wk) | 3/8 (-1 wk) | 1/8 (-1 wk) | 2/8 (-0.5 wk) |
| 50-64 | 1/8 (-1 wk) | 4/8 (+0.5 wk) | 2/8 (-1 wk) | 1/8 (-1 wk) | 2/8 (-0.5 wk) |
| 65+ | 3/8 (-0.5 wk) | 6/8 (+1 wk) | 3/8 (-1 wk) | 1/8 (-1 wk) | 4/8 (-0.5 wk) |

**Table S5. Season-level peak-timing analysis: fraction of seasons (of 8) in which children 5–17 years led the statewide total or the rest-of-state signal at peak week and half-maximum week, with median lead in weeks.**

| **Metric** | **Comparison** | **5-17 leads (fraction of seasons)** | **Median lead (weeks)** |
| --- | --- | --- | --- |
| Peak week | vs. statewide total | 1/8 | 0 |
| Peak week | vs. rest-of-state  (excl. 5-17) | 3/8 | 0 |
| Half-max week | vs. statewide total | 1/8 | 0 |
| Half-max week | vs. rest-of-state  (excl. 5-17) | 4/8 | 0.5 |

**Table S6. Rank of children 5–17 years' weekly activity (of 5 age groups) in the weeks leading up to the statewide peak, across 8 seasons.**

| **Weeks before statewide peak** | **5-17 ranked #1 (of 5 groups)** | **Mean rank (of 5)** | **n seasons** |
| --- | --- | --- | --- |
| 1 | 3 | 2.62 | 8 |
| 2 | 4 | 2 | 8 |
| 3 | 3 | 2.88 | 8 |
| 4 | 2 | 3.75 | 8 |
| 5 | 2 | 3.75 | 8 |
| 6 | 3 | 2.62 | 8 |

**Table S7. Sensitivity of the bivariate correlation between early-season children 5–17 activity and eventual peak magnitude to the early-season window length and onset threshold used to define it (Spearman's ρ).**

| **Analysis** | **Spearman rho** | **p-value** |
| --- | --- | --- |
| 2-week window, 3% onset threshold | 0.905 | 0.0046 |
| 2-week window, 5% onset threshold | 0.786 | 0.0279 |
| 2-week window, 7% onset threshold | 0.714 | 0.0576 |
| 2-week window, 10% onset threshold | 0.714 | 0.0576 |
| 3-week window, 3% onset threshold | 0.833 | 0.0154 |
| 3-week window, 5% onset threshold | 0.881 | 0.0072 |
| 3-week window, 7% onset threshold | 0.714 | 0.0576 |
| 3-week window, 10% onset threshold | 0.595 | 0.1323 |
| 4-week window, 3% onset threshold | 0.833 | 0.0154 |
| 4-week window, 5% onset threshold | 0.81 | 0.0218 |
| 4-week window, 7% onset threshold | 0.548 | 0.171 |
| 4-week window, 10% onset threshold | 0.524 | 0.1966 |
| 5-week window, 3% onset threshold | 0.857 | 0.0107 |
| 5-week window, 5% onset threshold | 0.81 | 0.0218 |
| 5-week window, 7% onset threshold | 0.548 | 0.171 |
| 5-week window, 10% onset threshold | 0.476 | 0.2431 |
| 6-week window, 3% onset threshold | 0.81 | 0.0218 |
| 6-week window, 5% onset threshold | 0.762 | 0.0368 |
| 6-week window, 7% onset threshold | 0.738 | 0.0458 |
| 6-week window, 10% onset threshold | 0.619 | 0.115 |
